# Decreased levels of soluble Developmental endothelial locus-1 are associated with thrombotic microangiopathy in pregnancy

**DOI:** 10.1101/2022.12.12.22283397

**Authors:** Gioulia Romanidou, Theocharis G. Konstantinidis, Konstantia Kantartzi, Maria Panopoulou, Emmanouil Kontomanolis, Christina Tsigalou, Maria Lambropoulou, Eleni Gavriilaki, Stylianos Panagoutsos, Ploumis Passadakis, Ioannis Mitroulis

## Abstract

HELLP syndrome is a life-threatening complication of pregnancy, which is often secondary to preeclampsia. To date, there is no biomarker in clinical use for the early stratification of women with preeclampsia that are under increased risk for HELLP syndrome. Herein, we show that the levels of circulating developmental endothelial locus-1 (DEL-1), which is an extracellular immunomodulatory protein, are decreased in patients with HELLP syndrome compared to preeclampsia. DEL-1 levels are also negatively correlated with the circulating levels of kidney injury molecule-1 (KIM-1), which is a biomarker for disorders associated with kidney damage. Receiver-operating characteristic curve analysis for DEL-1 levels and the DEL-1 to KIM-1 ratio demonstrated that these values could be used as a potential biomarker that distinguishes patients with HELLP syndrome and preeclampsia. Finally, we show that placental endothelial cells are a source for DEL-1 and the expression of this protein in placenta from patients with HELLP syndrome is minimal. Taken together, this study shows that DEL-1 is downregulated in HELLP syndrome both in the circulation and at the affected placental tissue, suggesting a potential role for this protein as a biomarker, which has to be further evaluated.

## INTRODUCTION

HELLP syndrome is a life-threatening complication in pregnancy characterized by hemolysis, elevated liver enzymes, low platelet and associated with increased maternal and neonatal morbidity and mortality [1]. It occurs in less than 1% of pregnancies [1] and it develops as a complication in 10-20% of women with preeclampsia [1]. Preeclampsia, presented as hypertension and proteinuria, is the result of abnormal placentation and endothelial dysfunction that affect several organs and can be complicated with liver damage and thrombotic microangiopathy (TMA), which are the features of HELLP syndrome [2].

Increased levels of soluble fms-like tyrosine kinase 1 (sFLT1), which is an anti-angiogenic protein, and decreased of the proangiogenic proteins placental growth factor (PlGF) are observed before the onset of pre-eclampsia and are used for the early diagnosis of the disorder [3, 4]. This deregulations in the balance of placenta-derived factor that affect the vasculature, such as sFLT1, PIGF, endoglin, as well as endothelin, is a key event in the pathogenesis of preeclampsia and HELLP syndrome [5]. For instance, induced expression sFlt-1 in pregnant rats has been shown to induce kidney injury [4]. The resulting endothelial cell dysfunction is coupled to inflammation due to activation of immune cell populations and complement system [6]. Importantly, complement activation has been proposed to mediate the thrombotic microangiopathy that characterizes HELLP [6, 7].

Kidney injury molecule-1 (KIM-1) is transmembrane glycoprotein that is upregulated in tubular cells upon kidney injury and its measurement in urine and blood can serve as a sensitive biomarker [8–10]. Since preeclampsia and, particularly, HELLP syndrome are associated with proteinuria and acute kidney injury, we measured KIM-1 levels in the circulation of such patients [2]. Additionally, we evaluated the levels of soluble developmental endothelial locus-1 (DEL-1), an extracellular matrix (ECM) protein produced and released by different cell populations, including endothelial cells and macrophages [11, 12], that regulates inflammatory responses via interactions with integrins and phospholipids [13].

Here, we shed light on the role of developmental endothelial locus-1 (DEL-1) on thrombotic microangiopathy in pregnancy. Moreover, we analyze the correlation of DEL-1 with new biomarker of renal function KIM-1. Finally, we analyze the usefulness of DEL-1 to KIM-1 ratio as a potential biomarker that distinguishes patients with HELLP syndrome and preeclampsia.

## MATERIALS AND METHODS

### Study Design

This study included 92 pregnant women (preeclampsia n=44, HELLP n=13, and control n=35), with mean age 32.38±1.59 yrs. The diagnosis of the preeclampsia and HELLP syndrome was based on the diagnostic criteria of the International Society for the Study of Hypertension in Pregnancy (ISSHP) [14]. For the diagnosis of HELLP syndrome the concomitant abnormal values of LDH, liver enzymes and platelet counts were necessary. Levels of ADAMTS13 were measured and anti-phospholipid screening was performed in all samples from patients with HELLP syndrome, to exclude thrombotic thrombopenic purpura and anti-phospholipid syndrome as possible alternative diagnosis for thrombotic microangiopathy. Serum and urine samples were collected on admission to the hospital and immediately stored at -80oC. Placental tissue from 2 patients with HELLP syndrome and 2 women with uncomplicated pregnancy were collected. The period of sampling was from June 2014 to May 2020. The study was performed at the University General Hospital of Alexandroupolis and Democritus University of Thrace in Alexandroupolis, Greece. The study was conducted according to the guidelines of the Declaration of Helsinki and approved by Institutional Ethics Review Board of the University Hospital of Alexandroupolis, Greece (Ethics Committee identification code: 941).

### Immunoassays

To estimate the renal function serum creatinine was measured using an automated biochemical analyzer, and the estimated glomerular filtration rate (e-GFR) was calculated using CKD EPI 2012 formula. Serum Kidney Injury Molecule 1 (KIM-1) was measured by ELISA, using commercially available kits in according to the manufacturer’s instructions (Cusabio, Wuhan, Hubei Province 430223, P.R.China). sFLT1/PIGF ratio was measured by the fully automated Elecsys® immunoassay on the electrochemiluminescence immunoassay platform (Roche Diagnostics, Mannheim, Germany). Human EGF-like repeat and discoidin I-like domain-containing protein 3 (EDIL3) or DEL-1 (Developmental endothelial locus 1) was measured by ELISA, using commercially available kits, according to the manufacturer’s instructions (Cusabio, Wuhan, Hubei Province 430223, P.R.China). ADAMTS13 activity, was determined by Technozym ELISA (Technoclone Herstellung von Diagnostika und Arzneimitteln GmbH 1230 Vienna, Austria)

### Immunohistochemical staining

Serial 4 mm sections of tissue blocks were obtained using a Leica RM2030 automated microtome (Leica Microsystems, Germany). Human placental tissue slides were incubated at 80° C for 30 min for deparaffinization and then in xylene solution for another 10 min. Descending ethanol solutions (100%, 96%, 70%, 50%) were used for tissue rehydration followed by a 15 min incubation with 0.3% H_2_O_2_ to quench endogenous peroxidase activity. All tissue samples were stained with the peroxidase method (Envision FLEX, Mouse/Rabbit detection System, High pH, DAKO, Carpinteria, CA, USA). Antigen retrieval was performed using EnVision FLEX Target Retrieval Solution High pH (50x) (Catalog No K8004). Slides were incubated with 1:100 diluted primary rabbit anti-DEL-1 antibody (Novus Biologicals, LLC, 10730 E. Briarwood Avenue, CO 80112, USA) at 4° C overnight. In parallel, control slides were incubated with non-immunized rabbit serum (negative control), while a positive control from human gastric carcinoma was always used. Visualization of the antibody antigen complex occurred after a 10-min incubation with the EnVision™ FLEX diaminobenzidine (DAB) chromogen (DM827, DAKO, Carpinteria, CA, USA). Sections were mounted and examined using a Nikon Eclipse 50i microscope (Nikon Instech CO. LTD., Kawasaki, Japan).

### Statistical analysis

Results are presented as mean±SEM. Data were analyzed using Kruskal-Wallis test followed by Dunn’s multiple comparison test. Correlation analysis was performed by Spearman r test. Statistical analyses were performed using GraphPad Prism software (GraphPad Inc., La Jolla, CA). Statistical significance is set at p< 0.05.

## RESULTS

The levels of soluble DEL-1 were quantified in the serum of control women with uncomplicated pregnancy (n=35), preeclampsia (n=44) or HELLP syndrome (n=13). Patient demographic and clinical characteristics are described in table 1. The levels of DEL-1 were decreased in the serum of patients with HELLP compared to the two other groups, whereas there was no difference between the control group and patients with preeclampsia (Figure 1A). On the other hand, there was no difference in the sFLT1/PIGF ratio between patients with preeclampsia and HELLP1, which is used as a marker for diagnosis of preeclampsia (Figure 1B). These findings suggest that the measurement of soluble DEL-1 levels could be used as a biomarker for the diagnosis of HELLP syndrome. To test the accuracy of DEL-1 measurement in the diagnosis of HELLP in the population of patients with hypertensive complications of pregnancy, we performed a receiver-operating characteristic (ROC) curve analysis. Area under curve (AUC) was 0.8187±0.064 (p=0.0006) and levels <676 pg/mL could be used for the diagnosis of HELLP with a specificity of 78.57% and a sensitivity of 76.92% (likelihood ratio=3.59) (Figure 1C).

**Table 1.** Clinical and demographic characteristics of study population

|  | Preeclampsia<br>*n*= 44 | HELLP<br>*n*= 13 | Controls<br>*n*= 35 |
| --- | --- | --- | --- |
| Age (years), mean (±SD), | 33.2±2.8 | 34.6 ±5.2 | 29.9±4.3 |
| Systolic BP,mean (±SD), mm Hg | 153.04±7.6 | 175.3±9.9 | 128.3±7.1 |
| Diastolic BP,mean (±SD), mm Hg | 88.6±3.4 | 96.7±8.05 | 74.7±7.5 |
| Gestational age (weeks) | 34.07±1.8 | 32.6±3.8 | 38.09±2.7 |
| Laboratory findings |  |  |  |
| WBC (±SD) K/μl | 13242.6±4915.3 | 11963.0±28635.3 | 11237.8±1477.8 |
| Hb (±SD) g/dL | 11.1±0.91 | 11.7±1.18 | 12.2±0.5 |
| Platelets (±SD) K/μl | 208.9±24.6 | 92.3±27.8 | 231.9±26.9 |
| MPV (±SD) | 11.12±0.78 | 11.6±1.04 | 8.8±0.82 |
| Glucose (±SD) mg/dL | 97.1±12.05 | 107.1±21.15 | 95.01±16.3 |
| eGFR | 112.5±12.3 | 110.±23.8 | 106.3±26.9 |
| Creatinine mean (±SD) mg/dL | 0.68±0.008 | 0.66±0.013 | 0.56±0.005 |
| Urea mean (±SD) mg/dL | 23.38±6.7 | 28.7±13.7 | 19.72±4.04 |
| SGOT (±SD) U/L | 22.8±8.1 | 240±98.4 | 16.4±2.3 |
| SGPT (±SD) U/L | 18.8±8.6 | 182.2±121.4 | 14.3±3.5 |
| Uric acid (±SD) mg/dL | 6.13±0.8 | 6.8±1.9 | 4.6±1.04 |
| CRP (±SD) mg/dL | 1.5±1.04 | 1.2±1.4 | 0.67±0.5 |

**Figure 1.**
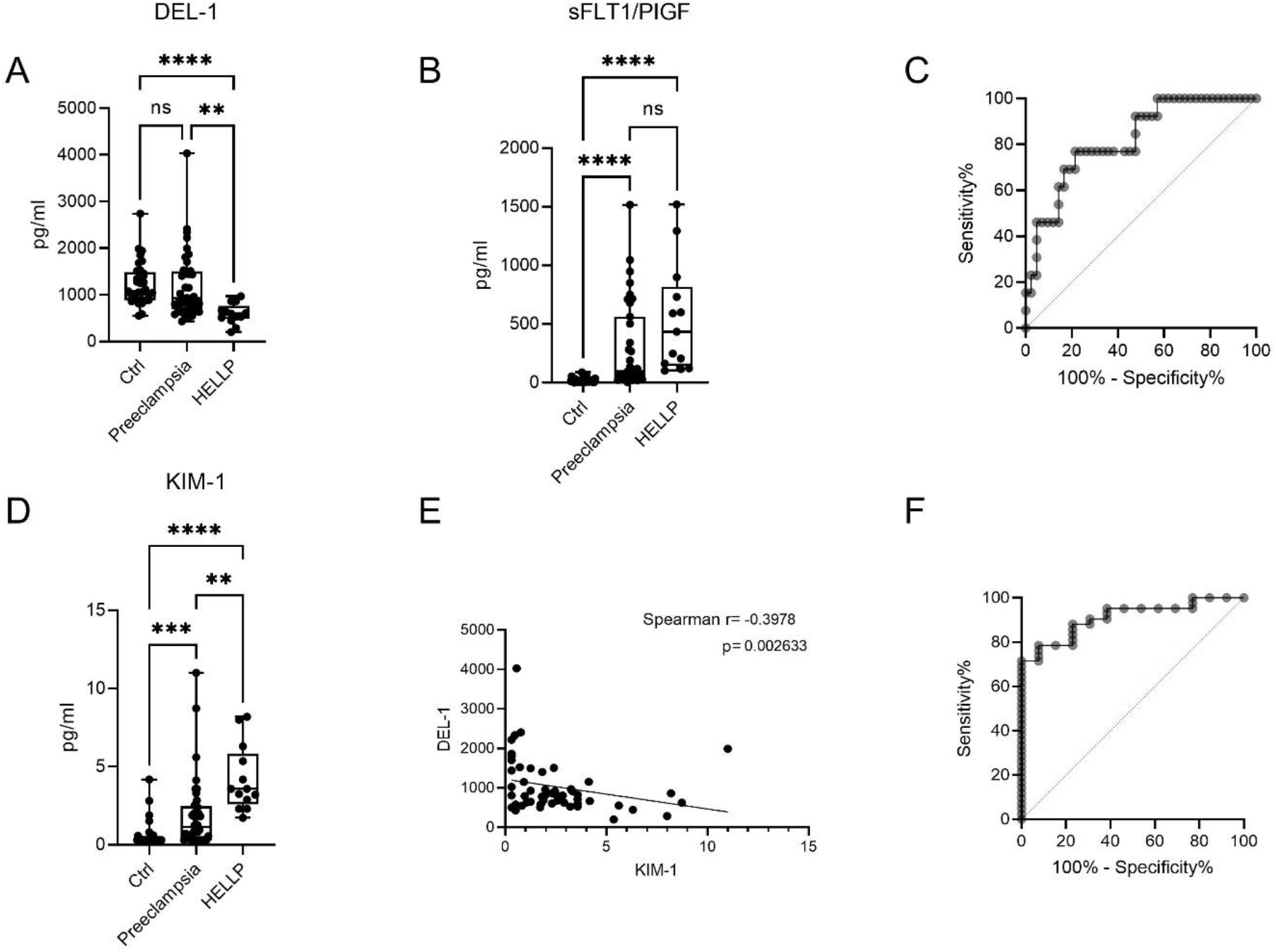
Soluble DEL-1 levels as a biomarker for the diagnosis of HELLP syndrome. A. DEL-1 levels in the serum of women with uncomplicated pregnancies (Ctrl), preeclampsia and HELLP syndrome. B. The ratio sFLT1/PIGF in the same group of patients. C. ROC analysis showing a significantly higher AUC for DEL-1 levels distinguishing HELLP syndrome from preeclampsia. D. KIM-1 levels in the serum and (E) correlation between DEL-1 levels and KIM-1 levels in patients with preeclampsia and HELLP syndrome. F. ROC analysis showing a significantly higher AUC for DEL-1 to KIM-1 ratio, distinguishing HELLP syndrome from preeclampsia. **p<0.01, ***p<0.001, ****p<0.0001.

We next measured the blood levels of KIM-1, a marker of kidney injury, and we observed that the levels of KIM-1 were increased from control subjects to patients with pre-eclampsia and then, to patients with HELLP (Figure 1D). The levels of KIM-1 were inversely correlated with those of DEL-1 (Figure 1E). Since there is an upregulation of KIM-1 levels and a downregulation of DEL-1 in HELLP syndrome compared to preeclampsia, we assessed whether the DEL-1 to KIM-1 ratio could be used to distinguish patients with HELLP syndrome from those with preeclampsia. ROC curve analysis revealed that the specificity and the sensitivity of values of DEL-1 to KIM-1 ratio < 263.1 were 88.1% and 76.92%, respectively (AUC=0.9103±0.039, p<0.0001, Likelihood ratio=6.462) (Figure 1F).

Endothelial cells are a major source of DEL-1 [11]. To test whether the aforementioned decrease in the circulating DEL-1 levels in HELLP syndrome is due decreased production by placental cell populations, we performed immunohistochemical analysis of placental tissue from control subjects and patients with HELLP. We observed that placental endothelial cells and perivascular were positive for DEL-1 in control tissue specimens (Figure 2A). However, endothelial cells in placenta from patients with HELLP syndrome did not express DEL-1 (Figure 2B). Taken together, the decreased circulating levels of DEL-1 can be attributed, at least in part, to the decreased expression of this protein by placental endothelial cells.

**Figure 2.**
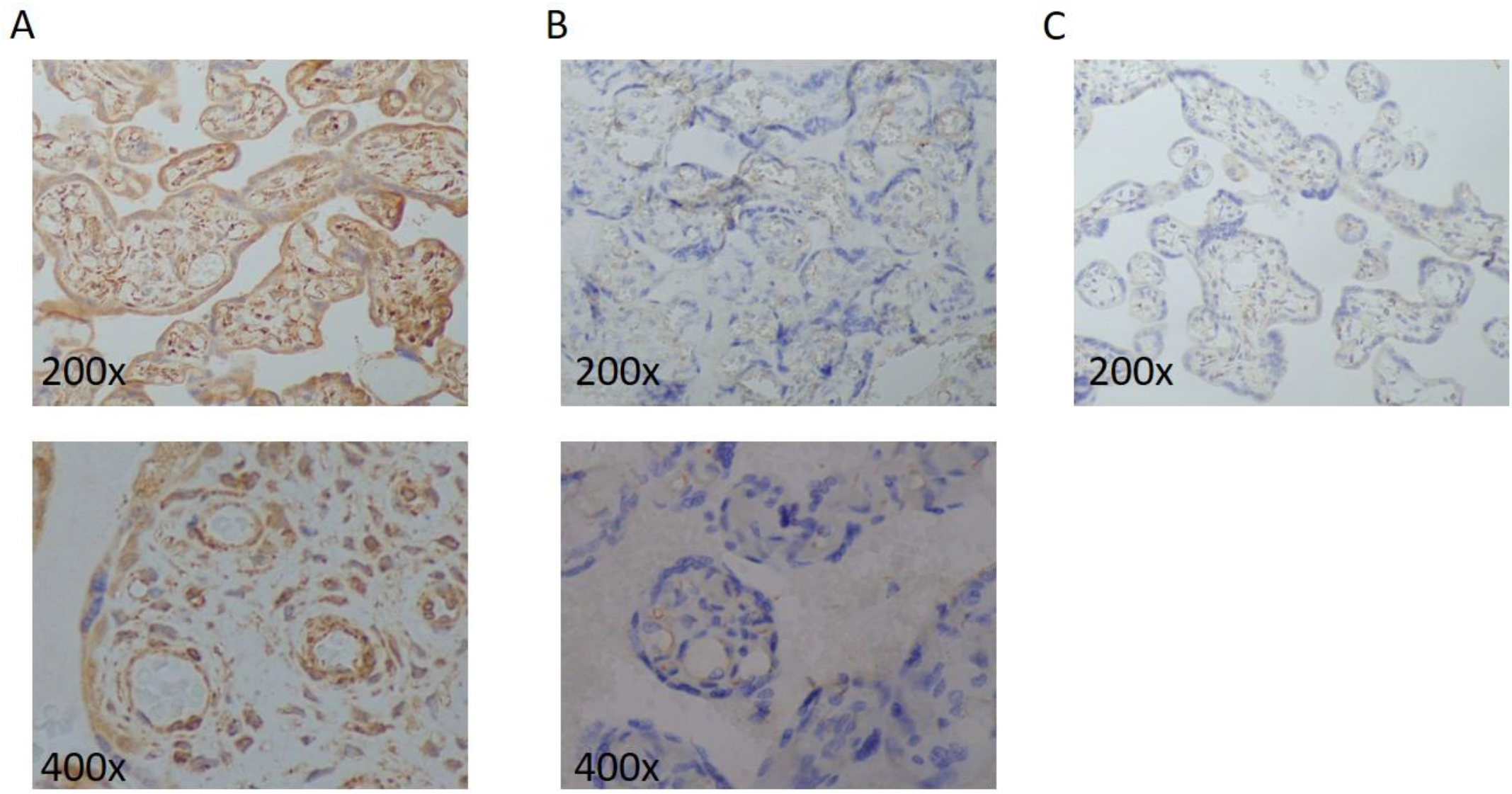
Expression of DEL-1 in placental tissue from patients with HELLP. A. Expression of DEL-1 in tissue sections from a woman with uncomplicated pregnancies assessed by immunohistochemistry and (B) from a patient with HELLP syndrome. Representative data from samples from two individuals per group. C. Technical negative control.

## DISCUSSION

HELLP syndrome is a life-threatening complication of pregnancy, which in the majority of cases develops secondary to pre-eclampsia [1]. Endothelial dysfunction and damage are the hallmark of the hypertensive complications of pregnancy [15]. Endothelial cell activation has been previously associated with increased levels of endothelial cell-derived factors in the circulation, such as thrombomodulin, E-selectin and von Willebrand factor [16, 17]. Herein, we show that the levels of another endothelial cell-derived factor, DEL-1, were decreased in the circulation of patients with preeclampsia compared to control, and further decreased in patients with HELLP syndrome. DEL-1 is a 52-kDa glycoprotein consisting of three *N*-terminal epidermal growth factor (EGF)-like repeats and two *C*-terminal discoidin I-like domains [13] that was initially proposed to mediate the adhesion of endothelial cells to ECM through interactions between the RGD motif that is present at the EGF-like repeat and αvβ3 integrin [18]. Further studies proposed that DEL-1 is an angiogenic protein [19] before the description of its role in leukocyte adhesion to endothelial cells [11]. Since then, several functions have been proposed for DEL-1, including inhibition of leukocyte recruitment in inflammatory disease models [20, 21] and cancer [22], clearance of apoptotic cells [12], induction of myelopoiesis [23] or support of complement-dependent phagocytosis [24]. Interestingly, it has been shown that DEL-1 restrains ischemia-induced neovascularization by modulating inflammation [25], whereas it had a protective role in the progression of hypertension and cardiac remodelling in a mouse model of angiotensin II-induced hypertension [26]. Herein, we show that DEL-1 is downregulated in the circulation and placenta from patients with HELLP syndrome. We show that placental endothelial cells are a source of DEL-1, even though we cannot exclude that the decreased levels of circulating DEL-1 could be attributed to endothelial cells and other cell types from different organs. Based on the multiple immunomodulatory functions of DEL-1 and the inflammatory nature of HELLP syndrome, we believe that this downregulation could play a significant, yet unknown role, in the pathogenesis of HELLP syndrome, affecting placental and/or systemic inflammation.

This decrease in the levels of DEL-1 was coupled with increased levels of KIM-1, a protein released upon kidney injury. This enabled the use of the DEL-1 to KIM-1 ration as a possible diagnostic tool for HELLP syndrome. Further prospective studies are needed to assess whether the measurement of these two biomarkers could be used for the early identification of pregnant women with preeclampsia, which are under increased risk for progression to HELLP syndrome.

Taken together, we propose that DEL-1 could be a potential prognostic biomarker for HELLP syndrome. Its role in the pathogenesis of the syndrome or its potential use as therapeutic target need to be further investigated.

## Data Availability

All data produced in the present study are available upon reasonable request to the authors

## Funding

EG and IM were supported by the General Secretariat for Research and Technology Management and Implementation Authority for Research, Technological Development and Innovation Actions (MIA-RTDI) (grant T2EDK-02288, MDS-TARGET).

## Notes

### Competing Interest Statement

The authors have declared no competing interest.

### Author Declarations

The study was conducted according to the guidelines of the Declaration of Helsinki and approved by Institutional Ethics Review Board of the University Hospital of Alexandroupolis, Greece (Ethics Committee identification code: 941).

